# Social mixing patterns relevant to infectious diseases spread by close contact in urban Blantyre, Malawi

**DOI:** 10.1101/2021.12.16.21267959

**Authors:** Deus Thindwa, Kondwani C Jambo, John Ojal, Peter MacPherson, Mphatso Dennis Phiri, McEwen Khundi, Lingstone Chiume, Katherine E Gallagher, Robert S Heyderman, Elizabeth L Corbett, Neil French, Stefan Flasche

## Abstract

**Introduction:** Understanding human mixing patterns relevant to infectious diseases spread through close contact is vital for modelling transmission dynamics and optimisation of disease control strategies. Mixing patterns in low-income countries like Malawi are not well understood.

**Methodology:** We conducted a social mixing survey in urban Blantyre, Malawi between April and July 2021 (between the 2nd and 3rd wave of COVID-19 infections). Participants living in densely-populated neighbourhoods were randomly sampled and, if they consented, reported their physical and non-physical contacts within and outside homes lasting at least 5 minutes during the previous day. Age-specific mixing rates were calculated, and a negative binomial mixed effects model was used to estimate determinants of contact behaviour.

**Results:** Of 1,201 individuals enrolled, 702 (58.5%) were female, the median age was 15 years (interquartile range [IQR] 5-32) and 127 (10.6%) were HIV-positive. On average, participants reported 10.3 contacts per day (range: 1-25). Mixing patterns were highly age-assortative, particularly those within the community and with skin-to-skin contact. Adults aged 20-49y reported the most contacts (median:11, IQR: 8-15) of all age groups; 38% (95%CI: 16-63) more than infants (median: 8, IQR: 5-10), who had the least contacts. Household contact frequency increased by 3% (95%CI 2-5) per additional household member. Unemployed participants had 15% (95%CI: 9-21) fewer contacts than other adults. Among long range (>30 meters away from home) contacts, secondary school children had the largest median contact distance from home (257m, IQR 78-761). HIV-positive status in adults >18 years-old was not associated with increased contact patterns (1%, 95%CI -9-12). During this period of relatively low COVID-19 incidence in Malawi, 301 (25.1%) individuals stated that they had limited their contact with others due to COVID-19 precautions; however, their reported contacts were not fewer (8%, 95%CI 1-13).

**Conclusion:** In urban Malawi, contact rates, are high and age-assortative, with little behavioural change due to either HIV-status or COVID-19 circulation. This highlights the limits of contact-restriction-based mitigation strategies in such settings and the need for pandemic preparedness to better understand how contact reductions can be enabled and motivated.

## Introduction

Globally, respiratory tract pathogens carry a substantial burden of morbidity and mortality at all ages ^1–3^, with incidence highest in low-and middle-income countries and in populations where human immunodeficiency virus (HIV) prevalence is high. For instance, an estimated 318,000 pneumococcal deaths occured in 2015 of which 23,300 were among HIV-positive individuals ^1^. In 2019, there were an estimated 1.5 million deaths from tuberculosis (TB), with HIV a major risk factor for the development of active TB and mortality ^2^. By November 2021, 5.2 million deaths from Coronavirus disease (COVID-19) have been reported globally ^4^, and with detrimental impact on heathcare delivery, particularly in low-income countries (LICs) ^5^.

Transmission of respiratory tract pathogens requires close contact with infectious respiratory droplets, secretions or inhalation, making understanding of human social mixing patterns an essential part of the design of effective disease control strategies ^6^. Social contact type, frequency, duration and place have shown to vary substantially between different settings ^7, 8^. Local built environment, population characteristics, and social activities such as traditional, political, religious and leisure events are likely to play important roles in explaining contact patterns variations between countries ^9–11^. However, relatively little is known about social mixing patterns in LICs ^8^, including how these are affected by age, occupation, household size, HIV status, and non-pharmaceutical interventions (NPIs) during COVID-19 pandemic.

Social mixing studies in middle (MICs) and high (HICs) income countries have shown that individuals in the same age groups tend to have higher contact rates than with other age groups (age-assortative mixing) ^7, 8^, yet not much is known about contact patterns in LICs in the era of fairly high urbanisation ^12–14^. Intergenerational mixing between younger children and adults was evident in Zimbabwe, reflecting parental or guardian roles played by adults ^13^, though age-sex mixing patterns are not well knowm ^12, 13^. Commonly, people tend to make high number of contacts within a short distance of their homes, with this being most pronounced for people living close to their usual place of work ^9, 15^. Where commuting long distances to work is common through mass transport, outbreak containment becomes more difficult due to greater ease of wide spatial spread of respiratory infections ^16^.

Physical distancing and lockdown NPIs to limit the number and geographical spread of close contacts have been used extensively during the COVID-19 pandemic, especially in HICs and MICs where relatively; mobility is usually high, per capita hospitalisation and mortality during the first wave was high, and lockdowns were strictly observed ^17–20^. In LICs, the impact of NPIs on social contacts is less clear, with added uncertainty on whether individual contacts are affected by COVID-19 vaccination status due to perceived attitude of being protected from severe COVID-19 ^21^. However, this is unlikely due to relatively very low current COVID-19 vaccination coverage in LICs ^22^.

Unlike in HICs, urban neighbourhoods in LICs are predominantly comprised of high-density informal settlements, relatively larger households (extended families), have low rates of formal employment, and often high HIV prevalence ^23, 24^. Contact patterns may then differ substantially having, for example, much higher rates within than outside households, and a higher ratio of skin-to-skin to verbal contacts ^7, 13^. Despite these anticipated differences, very few mixing studies are from Africa ^9, 10, 13, 14, 17, 25, 26^, and with only three from LICs ^12–14^. Also, understanding how contact patterns vary by HIV status is important to inform targeted interventions to mitigate risks both to HIV-poisitive individuals and to their social contacts.

The main objectives of this study in urban Malawi was to (1) estimate age-specific daily rates of social contacts, (2) investigate factors associated with social contact rates, (3) explore the influence of sex, household, community, HIV status and distance of contact event place in social contact patterns and (4) explore self-reported changes in social contact behaviour during the COVID-19 pandemic in the context of Malawi not having had a formal “lockdown” at any stage during the pandemic.

## Methods

### Study design

A cross-sectional study was undertaken in Ndirande, Blantyre, Malawi between April and July 2021. Ndirande mainly comprises high-density informal residential neighbourhoods, with minimal town-planning, building regulations or access to municipal services such as roads, electricity, water or refuse collection. Buildings tend to be single storey, built by unqualified local artisans using low-cost and sometimes locally-made materials like fired bricks. The road network is extremely limited, making access by motor vehicle impassable for most parts of the suburb.

Our sampling frame was based-on a previous Blantyre household TB prevalence survey, conducted in 2018-2019, where adult HIV status was ascertained using Government-approved rapid diagnostic tests ^27^. The TB prevalence survey mapped high density neighbourhoods in Blantyre city into 72 clusters where 12,865 adults from randomly selected households were recruited proportion to cluster size. Ndirande, the site for the present social mixing study, contributed 14 clusters and 2,626 adults to the TB prevalence survey.

The targeted sample size for the present social mixing study was ≥1,080 participants from 393 households across 14 clusters, of whom 490 were adults aged ≥18 years-old (y), and 190 were HIV-positive adults taking ART. A randomly sample (proportional to cluster size) of household members from Ndirande clusters of TB prevalence survey was taken, aiming to recruit equal number of participants across six age groups of <1y (infants), 1-4y (preschool children), 5-14y (primary school children), 15-19y (secondary school children), 20-49y (adults), and 50+y (older adults) to give adequate power of ≥80% to measure the number of social contacts with standard deviation of no more than 50% of the mean number of daily social contacts, and to detect the true differences in the mean contact rates between age groups if in excess of 30% ^10, 28^. Age groups were chosen based on transmission patterns of a typical respiratory infection ^7, 12, 29^.

A household was defined as a group of family members or unrelated individuals living in the same compound and sharing food from the same kitchen ^30^. Household size was split into <4, 4-6, >6 members during analysis based on interquantile range of measured household size in the study. Using the sampling frame of households in the 14 Ndirande clusters, handheld geographical positioning system (GPS, Garmin ETrex 30) devices were used to identify previously enrolled households. After visiting and validating each household, the household head aged ≥18 years was invited to participate, along with their <18 years-old household members. If a household or an adult household member who participated in the TB prevalence survey was not found after two visits, they were replaced by next household on the list. If a particular age-group had reached it’s required sample size, recruitment was restricted to households with at least one member in the age-groups that had not reached saturation.

### COVID-19 dynamics during field study

To date, Malawi has experienced three COVID-19 waves (May to August 2020, January to March 2021, and June to September 2021) ^31^. Our social mixing study was conducted after the end of the second wave (April-May 2021) and during the first half of the third wave (June-July 2021). During our study period, Malawi had initially implemented Level 1 COVID-19 policies, which included social distancing, recommended face mask wearing, rotational working, limited transit vehicles, self-quarantine for returning travellers, and resctrictions of gatherings to100 people (From April-May 2021). From June-July 2021, Level 3 COVID-19 policies were enacted, including: restricted movements, enforced facemask wearing, closing of non-essential businesses, enforced social distancing, reduced public transportation capacity, and 10 people restricted gatherings. Throughout the study period between April and July 2021, school closures were not implemented.

### Data collection

The study outcomes were reported physical (participant’s skin to skin touch with a contact) and non-physical contacts (participant’s two-way close verbal conversation lasting for ≥5 minutes and with ≥3 words exchanged with a contact) during the time period between waking up in the morning and going to bed in the evening ^10^. The household head was interviewed on household characteristics and composition on behalf of other household members, and also responded to their own individual demographic characteristics, travel history, COVID-19 behavioural change, and social mixing events, and those of younger (<7 years-old) household members. Older children mostly reported their contacts with help of an adult. A house was considered well-ventilated if it had open windows, doors and ventilation within the cooking area (Supplementary Questionnaire).

Survey interviews were conducted across all days of the week to ensure representation of weekdays and weekends. Repeated contacts with the same individual were recorded once, noting the frequency and cumulative time. Open data kit (ODK, Nafundi, Seattle, USA) was used to capture participant’s responses electronically, with an embedded electronic physical address locator (ePAL, Tripod Software, Salford, UK) for geolocating places of contacts ^24^.

### Characteristics and determinants of social contact events

We tabulated the number and distribution of participants and their contacts by age, and the proportion and distribution of contact events by the day of the week when contacts occurred and number of contacts per participant, respectively. The number and proportion of physical and non-physical contact events were also tabulated by contact duration, frequency, location, relationship of participant to contact, and sex of contact.

Social contact patterns were characterised by computing the mean number of contact events for a set of potential factors (age, sex, occupation, education, HIV, day of the week, COVID-19 restriction, and household size), with bootstrap confidence intervals based on 1000 bootstrap replicates. Factors associated with the mean number of mixing events at *p*<0.10 in univariable analysis were retained in multivariable analysis if they reduced the Akaike Information Criterion (AIC) ^32^. To investigate factors associated with contact rates, a negative binomial mixed model with household random intercept terms was constructed, and the ratio of the mean number of contact events in each category of the factor relative to the reference category was calculated ^10^.

### Age-specific social mixing rates

Age-stratified contact matrices were generated to investigate interactions between age groups. Age-specific contact rates through contact matrices were constructed from the mean number of daily contacts between participants and their contacts using the ‘socialmixr’ R package ^33^. Age-based contact matrices were estimated based on the ratio of the measured probability of a contact event between individuals based on age group to a null model of the probability of that contact event under an assumption of random mixing. Contact probabilities under the null model were determined by proportion of the population in each given age category in Ndirande ^24, 30^. Our analyses were weighted by days of the week as well as reciprocity in contact patterns such that the total number of contacts from age group *i* to *j* were equal to the total number of contacts from age group *j* to *i* (*m_ij_w_i_* = *m_ji_w_j_*), where the elements *m_ji_* make up a contact matrix and *w_i_* is the population size in age group *i*. Thus, *c_ij_* is the daily mean contact rate given as *c_ij_* = *m_ji_*/*w_i_*^10, 33^.

For each contact matrix, the assortativity index *Q* was computed. The *Q* index is defined a coefficient of degree between pairs of linked age groups and quantifies the weight of mixing between individuals of the same age groups. A *Q* close to 0 represents little dependence of mixing patterns on age while *Q* =1 implies exclusivity of contacts within age groups ^34^. Age-specific contact matrices were stratified by physical vs. non-physical contact, sex, within vs. outside household, within vs. outside Ndirande community, and adult HIV-positive vs. HIV-negative status, as well as affected vs. unaffected by COVID-19 and sex interations.

### Spatial distance distribution of contact events

We used longitude and latitude GPS coordinates to calculate the Euclidian distance in metres between participants’ houses and places of contact events. The inverse cumulative distance distributions for physical and non-physical contacts and for the stratification by age groups in relation to the distances away from homes were estimated ^35^. All analyses were conducted in R v4.1. Results can be reproduced using the data and code in the GitHub repository ^36^

### Ethics

Ethical approval was granted by the College of Medicine Research Ethics Committee, University of Malawi (P.01/21/3244), and the London School of Hygiene and Tropical Medicine Research Ethics Committee (#22913). Informed consent was obtained from each participant aged ≥16 years, or from the parent or guardian of each individual aged <6 years, or from each participant aged 6-15 years with addition informed assent.

## Results

### Study area, participants and their contacts

A total of 378 (85.9% of all who were approached) households participated in the survey from across 14 clusters in Ndirande, Blantyre City. Non-participation was mostly due to relocation, and with only 3% refusals. The proportion of study households within each cluster (relative to total census-counted cluster households) and across clusters varied between 7.9-19.7% and 4.0-10.0%, respectively. The most common number of rooms per house was four, of which two or three were most commonly used as bedrooms. The median household size was 6 members (interquartile [IQR] range 4-7), with 53 (14.2%) households reporting having at least one smoker (cigarette, marijuana or local cigar; chingambwe). Charcoal was predominantly used as source of energy in 366 (96.8%) households, of which 200 (52.9%) reported indoor cooking, and of which 23 (11.5%) were not well ventilated (Figure S1).

Overall, 1,201 participants were recruited, including 65 (5.4%) infants, 230 (19.2%) preschool children, 265 (22.1%) primary school children, 188 (15.7%) secondary school children, 301 (25.1%) adults and 152 (12.7%) older adults. Of enrolled participants, 702 (58.5%) were female, 138 (30.0% of adults) were unemployed, and the median age was 15 years (IQR 5-32). Among all adults aged 18 years or older, 201 (43.3%) had primary education, 18 (3.9%) had no formal education, and 127 (27.1%) self-reported HIV-positive status, all of whom were taking ART. In total, there were 12,540 reported contacts of whom 213 (1.7%) were infants, 1,161 (9.3%) preschool chldren, 3,239 (25.8%) primary school children, 2,165 (17.3%) secondary school children, 4,765 (38.0%) adults, and 997 (7.9%) older adults. The median age of contacts was 18 years (IQR 10-32). 9,341 (74.5%) contacts occurred during Monday to Friday. The average number of contacts per person was 10.43, with 25%, 50% and 75% of participants reporting at most 7, 10 and 13 contacts, respectively (Figure 1, Table 1, Table S1).

**Figure 1.**
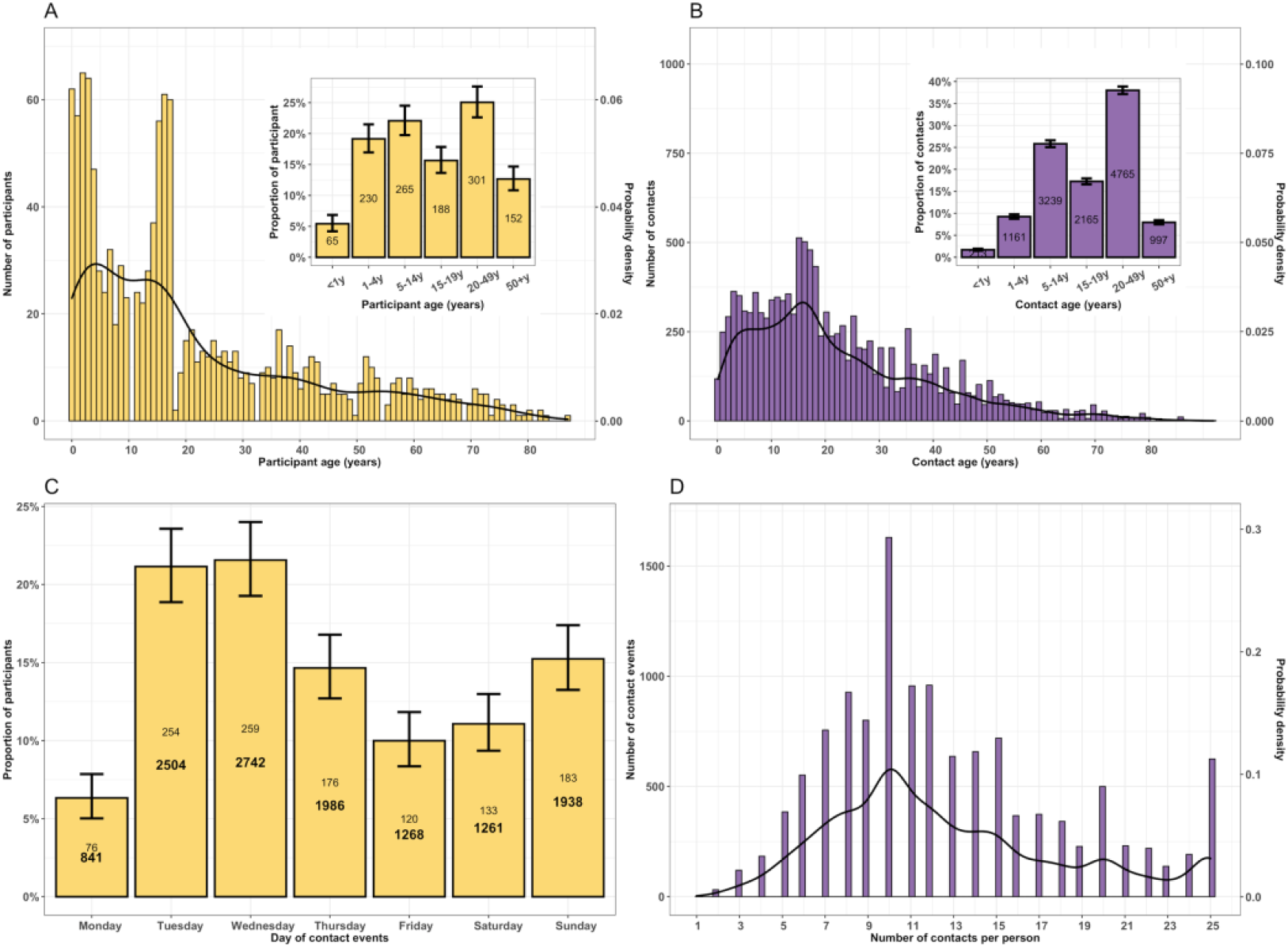
The probability distribution, number and proportion of study participants and reported contact events during the social contact patterns study in urban Blantyre, Malawi between April and July 2021. The age frequency and probability distribution of participants, with (A); proportion of participants in age groups of infants (<1 years-old), preschool (1-4 years-old), primary school (5-14 years-old), secondary school (15-19 years-old), adults (20-49 years-old) and older adults (50+ years-old) (insert); The age frequency and probability distribution of contacts, with proportion of contacts by age groups (insert); The number of contacts (bold fontface) and participants (roman fontface), and proportion of participants by day of the week when the contact event occurred (C); and the total number and probability distribution of reported contact events for a given number of contacts reported by the participant (D). Throughout the plots, the black line represents the probability density with values on secondary y-axis whereas yellow and purple colour represent participants and contacts, respectively. The 95% confidence interval for each bar is shown as vertical line.

**Table 1.** Characteristics of participants and their reported daily in Blantyre, Malawi, April-July 2021. The relative numbers of daily contacts (contacts rate ratio, CRR) were obtained from a negative binomial mixed model.

| Characteristic | Number of participants (%) | Mean number of contacts (95%CI) | Crude CRR (95%CI) | Adjusted CRR (95%CI) |
| --- | --- | --- | --- | --- |
| Age | (N = 1,201) |  |  |  |
| <1y | 65 (5.4) | 8.14 (7.26-9.15) | Ref | Ref |
| 1-4y | 230 (19.1) | 8.63 (8.18-9.08) | 1.14 (1.03-1.25) | **1.13 (1.03-1.25)** |
| 5-14y | 265 (22.0) | 10.24 (9.72-10.82) | 1.29 (1.17-1.42) | **1.23 (1.07-1.41)** |
| 15-19y | 188 (15.7) | 10.88 (10.18-11.61) | 1.40 (1.27-1.54) | **1.31 (1.14-1.50)** |
| 20-49y | 301 (25.1) | 11.75 (11.17-12.38) | 1.50 (1.37-1.63) | **1.38 (1.16-1.63)** |
| 50+y | 152 (12.7) | 10.18 (9.43-11.15) | 1.38 (1.24-1.53) | **1.30 (1.08-1.56)** |
| Sex |  |  |  |  |
| Female | 702 (58.5) | 10.45 (10.08-10.8) | Ref | Ref |
| Male | 499 (41.5) | 10.06 (9.64-10.49) | 0.94 (0.90-0.97) | 0.97 (0.94-1.01) |
| Occupation |  |  |  |  |
| Business<sup>†1</sup> | 206 (17.2) | 12.13 (11.31-12.95) | Ref | Ref |
| Workers<sup>‡2</sup> | 79 (6.6) | 11.27 (10.13-12.63) | 0.96 (0.89-1.05) | 1.00 (0.92-1.08) |
| Pre-school child | 267 (22.2) | 8.74 (8.31-9.16) | 0.70 (0.66-0.74) | 0.90 (0.77-1.05) |
| Schoolers | 479 (39.9) | 10.28 (9.87-10.76) | 0.86 (0.82-0.91) | 0.95 (0.85-1.06) |
| Retired | 28 (2.3) | 8.79 (7.26-10.50) | 0.84 (0.73-0.97) | 0.91 (0.78-1.05) |
| Unemployed | 138 (11.5) | 10.34 (9.68-11.08) | 0.85 (0.79-0.91) | **0.85 (0.79-0.91)** |
| Other | 4 (0.33) | 9.50 (6.00-15.25) | 0.88 (0.62-1.25) | 0.91 (0.64-1.28) |
| Education level† |  |  |  |  |
| University/college | 17 (3.7) | 9.76 (7.35-12.23) | Ref | - |
| No education | 18 (3.9) | 11.00 (8.94-14.24) | 1.03 (0.76-1.40) | - |
| Primary school | 201 (43.3) | 11.48 (10.66-12.24) | 1.04 (0.83-1.32) | - |
| Secondary school | 228 (49.1) | 11.22 (10.54-11.99) | 1.03 (0.82-1.29) | - |
| HIV status‡ |  |  |  |  |
| Negative | 340 (72.8) | 11.28 (10.70-11.88) | Ref | - |
| Positive | 127 (27.1) | 11.13 (10.26-12.17) | 1.01 (0.91-1.12) | - |
| Interview day |  |  |  |  |
| Weekday (Monday-Friday) | 885 (73.7) | 10.43 (10.12-10.78) | Ref | - |
| Weekend (Saturday-Sunday) | 316 (26.3) | 9.90 (9.27-10.45) | 0.99 (0.92-1.06) | - |
| COVID19 restrictions* |  |  |  |  |
| No | 900 (74.9) | 10.04 (9.74-10.40) | Ref | Ref |
| Yes | 301 (25.1) | 11.03 (10.50-11.55) | 1.21 (1.16-1.27) | **1.08 (1.01-1.13)** |
| Household size |  |  |  |  |
| 1-3 people | 90 (7.5) | 8.81 (7.70-10.07) | Ref | Ref |
| 4-5 people | 305 (25.3) | 10.40 (9.69-11.24) | 1.20 (1.04-1.39) | **1.24 (1.07-1.43)** |
| 6 people | 230 (19.2) | 10.13 (9.50-10.84) | 1.21 (1.03-1.42) | **1.26 (1.07-1.47)** |
| 7+ people | 576 (48.0) | 10.80 (10.43-11.19) | 1.29 (1.12-1.49) | **1.36 (1.18-1.57)** |
| ‡1 Business refers to individuals involved in the exchange of goods and services to earn profits usually struggling self-employed traders<br>‡2 Workers include all persons involved in agricultural, domestic, manual, and office activities<br>† Only applicable to adults 18 years and older<br>95%CI - 95% Confidence Intervals<br>ART Antiretroviral therapy<br>* Whether or not individual reported that they had modified their social mixing behaviour due to the COVID-19 pandemic |  |  |  |  |

### Travel history and COVID19 impact

Approximately one quarter of participants (n=281, 23.4%) had travelled more than 5km outside of Ndirande community in the last 24 hours. However, of 1,041 (86.7%) participants who travelled outside Ndirande anytime in the past, 81 (7.8%) reported that they made similar travel daily and 161 (15.5%) at least once a week; 840 (80.7%) spent at most 24 hours outside Ndirande, and 913 (87.7%) used public transport during similar travels.

A total of 301 (25.1%) participants, almost exclusively adults, reported to have reduced their social contacts due to COVID-19 pandemic, with home (n=225, 74.8%) and market (n=183, 60.8%) being the main localities where contact was reportedly reduced. Among all participants, we estimated that preventive measures against the COVID-19 pandemic contributed 9.1% (95%CI 0-13) to social contacts reduction e.g. from the median of 11 (IQR 7-18) daily contacts, actual and hypothesised (contacts that could have occured in absence of COVID-19 pandemic), to 10 (IQR 7-13) actual contacts. However, actual contacts of those affected by COVID-19 were 8% (95% CI: 1-13) higher than their counterparts who reported no COVID-19 reduced social contacts (Table 1, Figure S2, Figure S3).

### Characteristics and determinants of reported mixing events

About 74% (8,266/1,1175) of contacts longer than 15 minutes involved physical contact whereas 60% (547/1,365) of shorter contacts did not. Contacts were likely physical if they occurred on a daily basis (7,648/10,058) compared to less frequent contacts. More than 80% (4,681/5,626) of physical contacts were with family members and only 30% (487/1,554) with unknown (random) people. About 75% (8,183/10,966) of contacts at school, home, leisure or in transport were physical whereas less than 50% (552/1,409) of contacts at church, market or work were. More than 95% (12,136/12,540) of all contacts happened within than outside the community (Figure 2).

**Figure 2.**
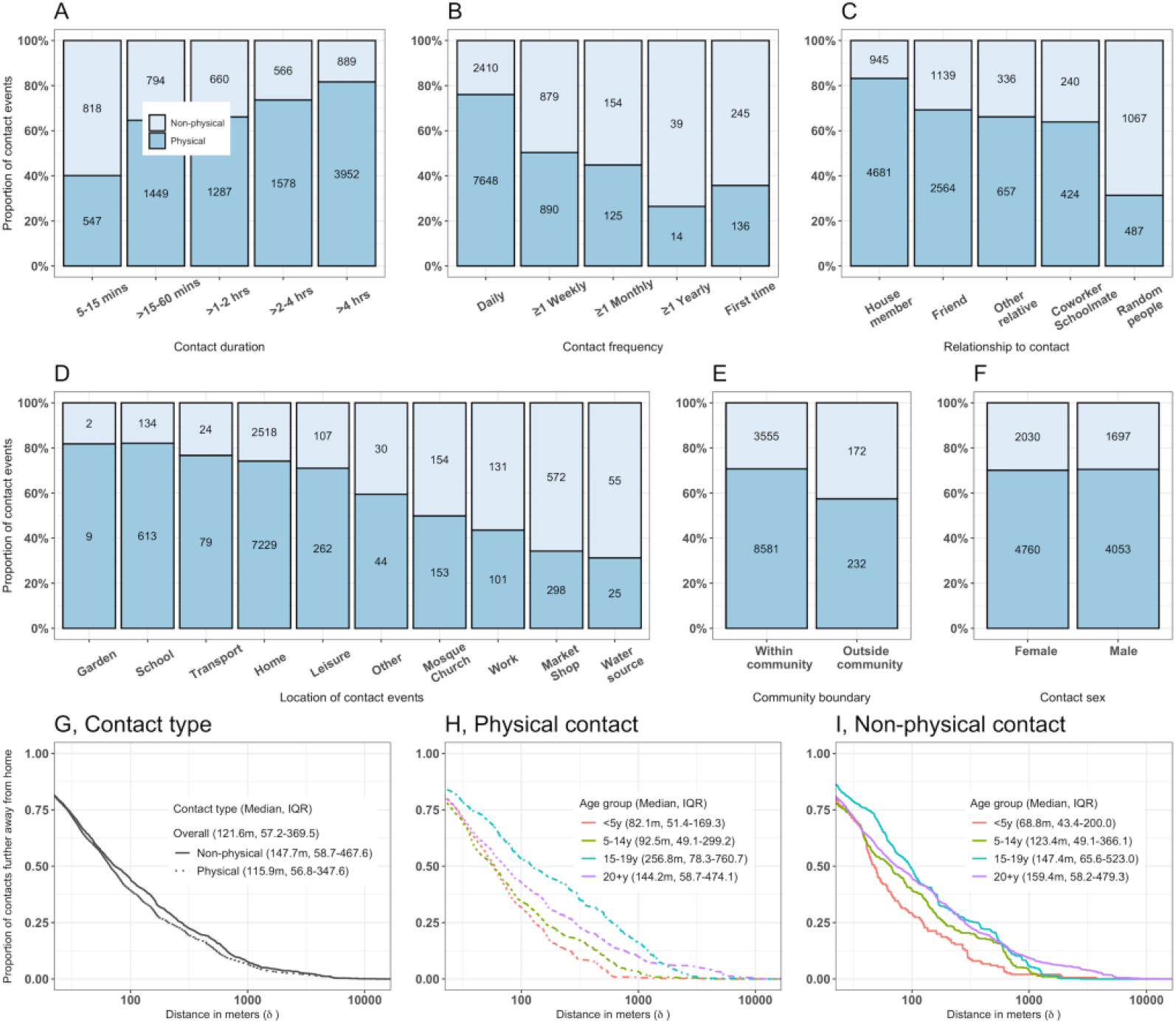
Characteristics of reported physical and non-physical social contacts in urban Blantyre, Malawi between April and July 2021. The proportion of physical and non-physical contacts by the duration of contacts (A), frequency of contacts (B), relationship of participant to contacts (C), the exact location of contacts (D), contacts within or outside community (E) and contact sex (F). The number in each bar plot indicates absolute number of reported contacts. Physical contact refers to participant’s skin to skin touch with a contact whereas non-physical contact refers to participant’s two-way close verbal conversation lasting for ≥5 minutes and with ≥3 words exchanged. The inverse cumulative distance distribution showing the proportion of contacts in relation to the distance (in metres) further away from the participant home by physical and non-physical contact type (G), for physical contact events by age group (H), and for non-physical contact events by age group (I).

Data on spatial distances between participant house and place of mixing was available for 12,449 (99.3%) social mixing events. Overall, the median Euclidian distance meters (m) away from home to where mixing events occurred was 121.6m (IQR 57.2-369.5), ranging from 30.0m to 12,358.8m. Secondary school children travelled furthest on average for their physical contacts (256.8m, IQR 78.3-760.7), primary school children had the most localised physical contacts (82.1m, IQR 51.4-169.3). (Figure 2).

The mean number of contacts varied significantly by age, with infants and preschool children reporting lower contacts (95%CI: 7.26-9.15) than those in older age groups (95%CI: 9.72-12.38). Similarly, households with at most three members had significantly lower contacts (95%CI: 7.70-10.07) than those with at least seven members (95%CI: 10.43-11.19). On the contrary, the mean number of contacts did not significantly differ by sex, education status, HIV status or interview day (Table 1).

In a multivariable analysis, age, occupation status, and household size were significantly associated with daily number of contacts. Adults had the highest contact frequency among all ages, over 30% more than infants (1.38, 95%CI 1.16-1.63). Unemployed adults (less active community members) had 15% (9-21) fewer contacts than business adults, usually self-employed traders (more active counterparts). Household contact frequency increased by 3% (95%CI 2-5) per additional household member. Members of households of at least 7 members had 36% (95%CI: 18-57) more contacts than participants from households with 1-3 members (Table 1).

### Age-specific mixing patterns

Social contacts were highly age-assortative, with the most intense contacts clustered among primary school children and secondary school children compared to infants, preschool children or adults. Non-physical contacts were less age-assortative (*Q* =0.076) than physical contacts (*Q* =0.118) and were mostly reported among adults, whereas children mostly reported physical contacts (Figure 3).

**Figure 3.**
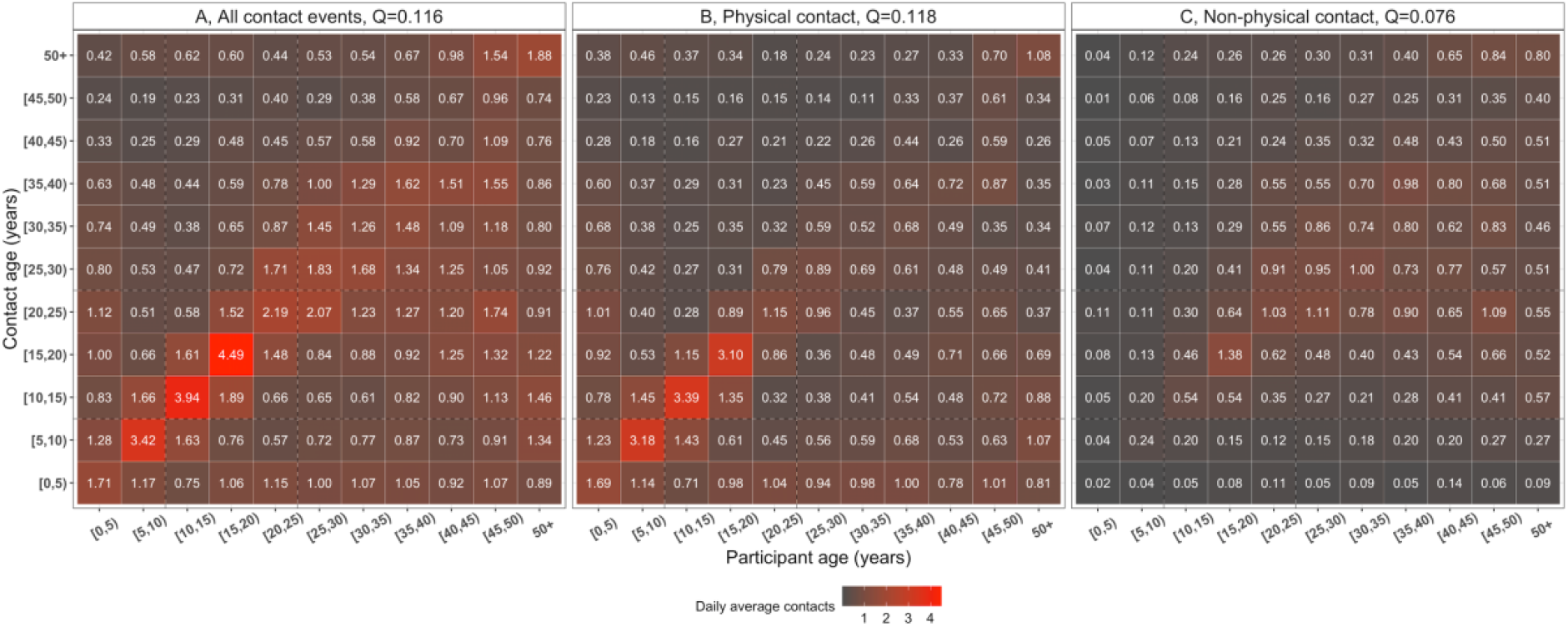
The daily mean number of reported contacts between age groups, in urban Blantyre, Malawi between April and July 2021. The number in each cell represents the daily mean number of contacts between two age groups from 1000 bootstrap replicates, corrected for reciprocity between participants and contacts, and weighted for day of the week. The matrices show the daily mean number of all contacts (A), physical contacts (participant’s skin to skin touch with a contact) (B) and non-physical contacts (participant’s two-way close verbal conversation lasting for ≥5 minutes and with ≥3 words exchanged with a contact) (C).

Age-assortativity was particularly pronounced for male-gender mixing patterns (*Q* =0.142), with women being more involved in intergenerationally mixing (*Q* =0.103). Participant-contact mixing between male-male (*Q* =0.420) was more intense and highly assortative than female-female (*Q* =0.163) or male-female (*Q* =0.106) or female-male (*Q* =0.096). Mixing outside household (*Q* =0.266) and Ndirande community (*Q* =0.196), was more age-assortative than within household (*Q* =0.073) and community (*Q* =0.113). No differential age-assortativity was seen on social contacts behaviour between HV-positive adults taking ART (*Q* =0.056) and HIV-negative adults (*Q* =0.071) (Figure 4, Figure S4, Figure S5).

**Figure 4.**
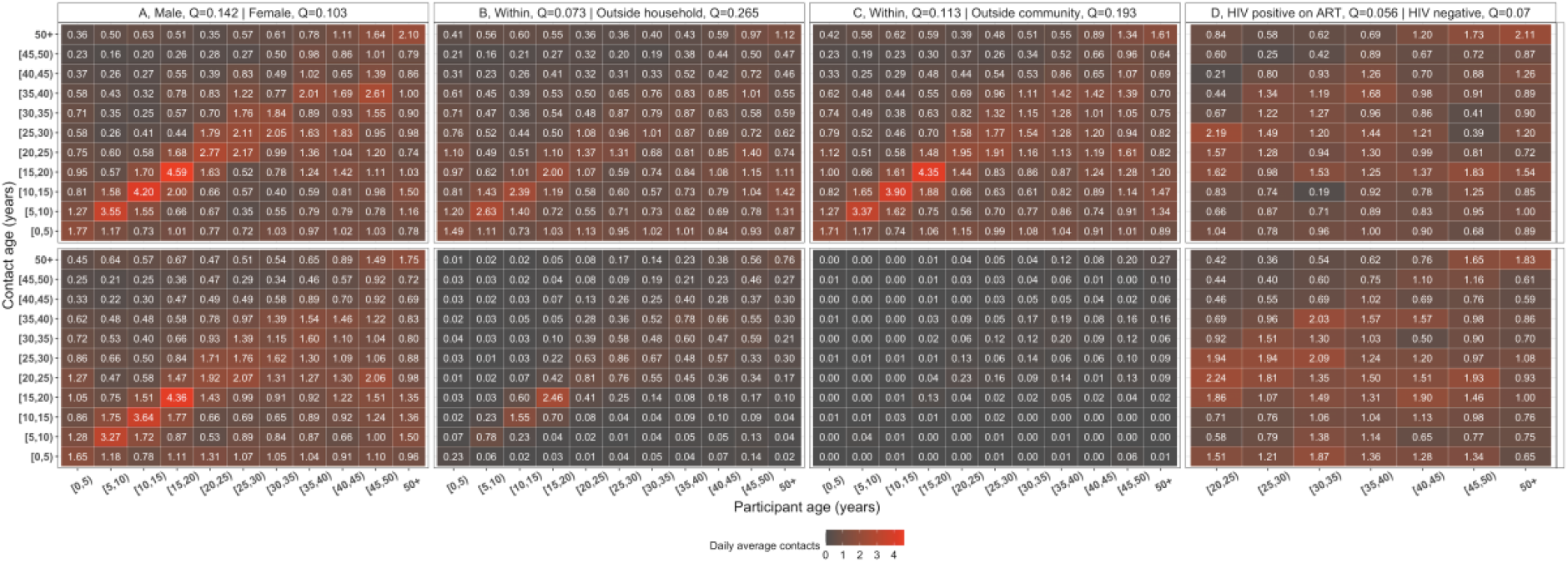
Daily mean number of reported contacts between age groups, in urban Blantyre, Malawi between April and July 2021. The number in each cell represents the daily mean number of contacts between two age groups from 1000 bootstrap replicates, corrected for reciprocity between participants and contacts, and weighted for day of the week. The top and bottom matrices show the daily mean number of contacts respectively stratified by male and female sex (A), within and outside household contacts (B), within and outside community contacts (C), Human Immunodeficiency Virus (HIV) positive adults (20+ years old) and HIV-negative adults (D). The assortativity index *Q* quantifies the weight of mixing between individuals of the same age groups and is estimated on the square matrix. For the HIV-stratified matrices, the index *Q* is estimated on the square matrix between pairs of adult participants and adult contacts aged at least 20 years old.

## Discussion

Surveying 1,201 individuals in high-density informal urban suburb in Malawi, the median number of contacts was higher than previously reported in Zambia ^12^, but similar to Zimbabwe and Somaliland and many MICs and HICs ^7, 13, 14^. Older age, employment, and bigger households were significantly associated with an increased number of contacts in this setting, consistent with previous social contacts studies ^7, 8^. Reported social contacts were strongly age-assortative and intergenerational by sex, mostly physical and occuring within household and geographically localised. Notably, neither HIV status, ART use nor COVID-19 pandemic restrictions significantly reduced population-level social contacts behaviour.

The reported median number of contacts of 10 is similar to reported contacts among internally diplaced people in Digaale camp in Somaliland ^14^, but higher than the median of 4 contacts reported in a LIC in Zambia ^12^, and lower than the 24 reported in Thailand ^37^ or 18 in rural Kenya ^25^, highlighting the substantial role local context play in social contacts. The completeness of contacts capture was strengthened by conducting participant’s interview only on second visit with advanced notice to remember contacts on the day, guided interview process collecting only contacts initials to start with and aiding participant’s memory by structuring the interview to prompt participants to the different typical parts of the day. Unlike MICs and HICs where the number of daily contacts usually are highest among school age children ^25, 38^, our finding that working age adults had the largest number of increasing daily contacts aligns with results from LICs in Zimbabwe and Somaliland ^13, 14^.

This suggests that schools may play a less prominent role in the transmission of infectious diseases in LICs than they do in other parts of the world; albeit that this may be counter-balanced by children comprising a generally larger share of the population in LICs.

The 15% relative higher average number of contacts among workers (often self-employed traders in this setting) than among unemployed individuals is in line with evidence from several previous studies irrespective of location ^7^, and reflects the influence of human mobility and trading activities on social contacts ^16^, which in part has motivated lockdowns in some MICs and HICs during the COVID-19 pandemic ^39^. Increasing number of contacts with house size has also been reported in some MICs and HICs ^7^, with household density suggested to be a driving factor particularly in this setting where extended families sharing a compound is common. We also report higher proportion of home contacts similar to other LICs and MICs than those reported in HICs, and by contrast, the proportion of school and work contacts were substantially lower than those in HICs ^7^. This implies that household may be a key site of transmission for respiratory diseases ^40^, and public preventive measures may only be efficacious at reducing the speed for spatial spread outside homes. However, the relevance of contact location on transmission will also depend on, among other things, specific pathogen and transmission routes e.g. droplet, fomite or aerosol ^7^.

The strong age-assortative contacts in this study are consistent with widespread evidence globally ^7, 8^, and support adjusting for age-heterogeneity when calibrating models for predictions ^41, 42^. Approximately 80% of contacts in this study were skin-to-skin, falling on the higher side of the reported range of 19-84% globally ^7^. While the predominance of physical contacts in children compared to adults is similar across all settings, substantial physical contacts in our study align with reports in LICs and MICs but not HICs ^7^. Social interactions by women were less strongly age-assortative than for men, consistent with a greater role for for intergenerational mixing between mothers or female guardians with younger children, as also reported in Zimbabwe and Kenya ^13, 25^. Age-sex interactions showed relatively high age-assortativeness among male-male contacts than other sex combinations, implying that relatively small underlying biological or behavioural difference in susceptibility by sex may be amplified by assortative social networks, as hypothesised for TB ^12^. Moreover, relatively high (97%) frequency of contacts occurring within rather than outside community in this study may imply that epidemics would mostly be localised. However, adults >15 years-old reported the highest intense physical contacts including minibus travel with potential to disseminate outbreaks outside of the local community.

Ours is the first study to evaluate whether HIV-positive status modulates contact behavior. In this high HIV prevalence population with good access to ART, we found no evidence that HIV status *per se* influences contact rates or age-assortativeness. Given that immunosuppressed individuals may be at increased risk of prolonged COVID-19 infectiousness and generation of new variants as well as increased pneumococcal carriage ^43, 44^, this suggests that HIV-positive individuals may have equivalent or increased potential for contributing to respiratory disease transmission in this setting ^45^. We have not, however, assessed the other drivers of pathogen transmissibility, such as carriage density and duration of infection, and so cannot confirm a likely disproportionate role in transmission.

Contrary to MICs and HICs who reported substantial reduction in social contacts due to NPIs ^17, 18, 20^, only 25% of participants in our study reported to have changes their contact behavior between the second and third covid wave in Malawi. Furthermore, we did not find reduced contact rates among participants who reported to have reduced their social contacts due to COVID-19 pandemic restrictions compared to those who reported to not have changed their behavior. This may suggest sub-optimal adherence to COVID-19 prevention measures such as restricted movements, closing of non-essential businesses, enforced social distancing, reduced public transportation capacity, and limited numbers (recommended 10 people per gathering) in this setting, which may reflect the day-to-day demands in the context of pressing economic hardship ^46–48^. Economic implications of policy options aimed at limiting transmission of respiratory diseases need to be considered in order to make pragmatic recommendations that can be adhered to in the local context. Of note, relatively few *per capita* hospitalisations and deaths from COVID-19 occurred during the first wave in Malawi despite evidence of widespread transmission, and this may explain low compliance with recommendations to reduce social contacts during the second and third waves of COVID-19 pandemic in urban Malawi.

The strengths of this study include a sufficiently large sample size to detect significant differences in contact rates between age groups, although small numbers limited our precision for infants. We sampled households from 14 clusters making up a high proportion of residential Blantyre. Substantial number of participants were recruited during weekdays (75%) and weekends for representation and contact matrices were weighted for weekdays and weekends and accounted for reciprocity. This study collected novel data on social contacts in Malawi, adding insights to limited primary datasets on contact patterns in LICs in Africa, with only 3 countries represented. Limitations include that reported contacts by participants may be subject to recall bias ^49^. This bias was partially addressed, however, by conducting two visits within three days; asking participants on the first visit to remember all their contacts during waking up and going to bed, and on the second visit, asking participants to report those contacts. Prospective reporting of cases reduces bias, but can lead to overreporting ^50^. Data on contacts within schools and other indoor spaces were not collected at fine scale, hence are difficult to assess. Social contacts may change over time ^19, 20^, and our cross-sectional design did not include longitudinal sampling of mixing patterns. Our results may not be generalisable to rural settings of Malawi that have relatively low household density and socio-economic activities but large school sizes compared to urban communities ^51^. Thus, it remains uncertain whether or not contacts would be low given mixed results from rural Zimbabwe and Kenya ^13, 25^.

In conclusion, high rates of physical, age-assortative and localised contacts were observed, particularly among secondary school children and adults. With the demographic shift that many LICs are undergoing this raises the potential for adults in this and similar settings to play a more prominent role in the transmission of respiratory diseases than typically the case in HICs. In addition, the lack of change in contact behavior in response to the ongoing pandemic highlights specific challenges for mitigation strategies in poor communities with no social protection mechanisms. For pandemic planning, it will be crucial to better understand what factors would enable and encourage poor urban populations to reduce their contacts to slow pandemic spread if the need arises.

## Data Availability

All data produced are available online at: https://github.com/deusthindwa/social.contact.rates.estimation.hiv.malawi

## Acknowledgements

We would like to thank Dr Emanuele Del Fava of the Carlo F. Dondena Centre for Research on Social Dynamics and Public Policy, Bocconi University, Milan, Italy and Max Planck Institute for Demographic Research, Rostock, Germany, and Kevin van Zandvoort of the London School of Hygiene and Tropical Medicine, in London, United Kingdom, for insightful discussions at the analysis stage of this work. We thank all Social Mixing Patterns (SOMIPA) participants for their participation in the study as well field workers for administering consents and collecting data. We also thank the Blantyre Directorate of Health and Social Services as well as the Blantyre City Council for their support to the study activities.

## Role of the funding source

This manuscript was funded by the National Institute for Health Research (NIHR) Global. Health Research Unit on Mucosal Pathogens using UK aid from the UK Government (16/136/46). D Thindwa, J Ojal, K E Gallagher, Robert S Heyderman, N French, and S Flasche are supported by the National Institute for Health Research (NIHR) Global Health Research Unit on Mucosal Pathogens using UK aid from the UK Government. RSH is a NIHR Senior Investigator. S Flasche is also supported by a Sir Henry Dale Fellowship jointly funded by the Wellcome Trust and the Royal Society (Grant number 208812/Z/17/Z). The views expressed in this publication are those of the author(s) and not necssarily those of the NIHR or the Department of Health and Social Care. The funders had no role in study design, data collection and analysis, decision to publish, or preparation of the manuscript.

## Declaration of interest

None

## Author contribution

Conceptualization; DT, SF, NF

Data curation; DT

Formal analysis; DT

Funding acquisition; DT, RSH

Investigation; DT

Methodology; DT, SF

Project administration; DT, KJ

Resources; DT, RSH

Software; DT

Supervision; SF, NF, KJ

Validation; DT, KCJ, JO, PM, MDP, MK, LC, KEG, RSH, ELC, NF, SF

Visualization; DT

Roles/Writing – original draft; DT

Writing -review & editing; DT, KCJ, JO, PM, MDP, MK, LC, KEG, RSH, ELC, NF, SF

All authors read and approved the final manuscript.

---

**Supplementary Figure 1.**
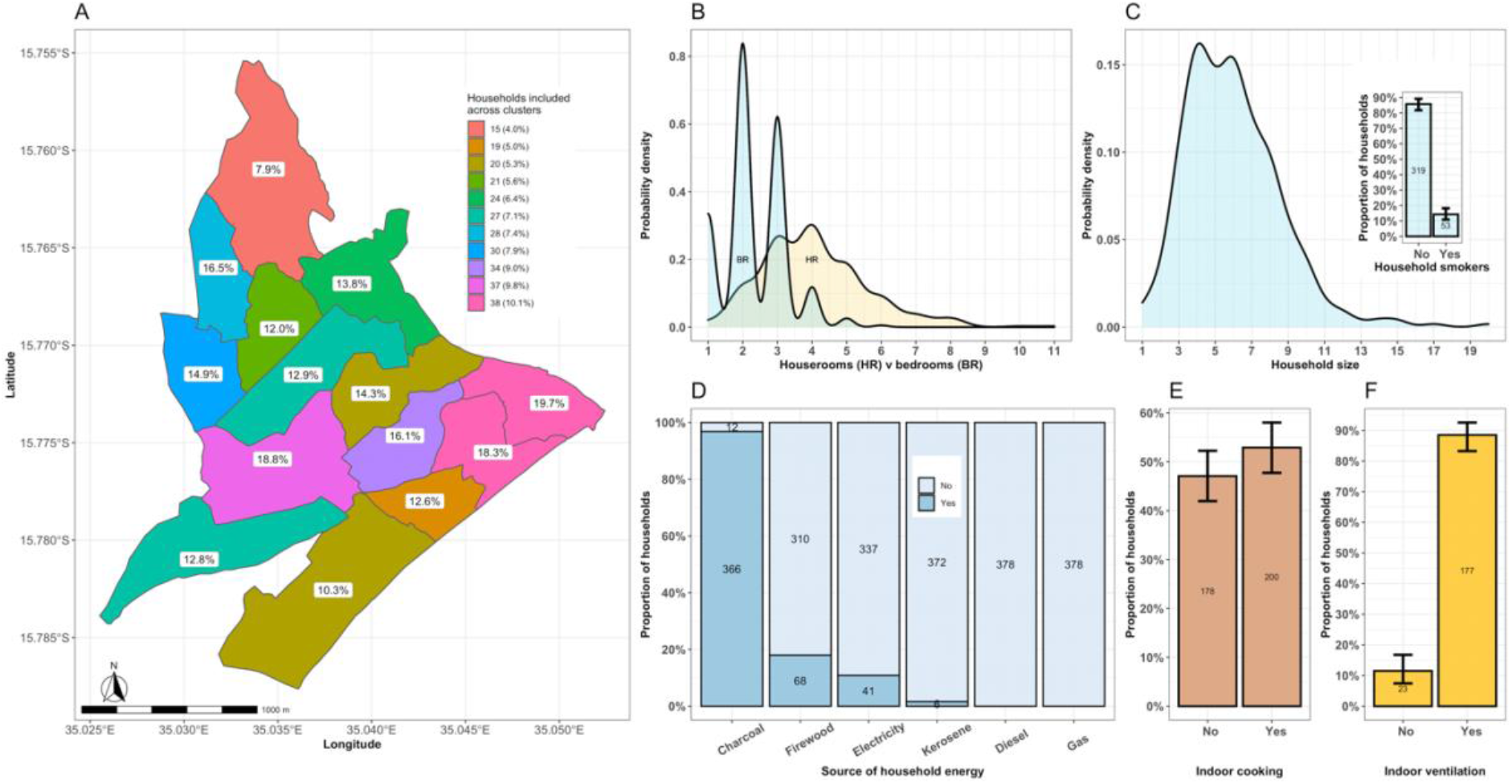
Community and household composition in urban setting of Blantyre, Malawi 2021. Map of sampled study households by cluster, where the absolute number and proportion in the legend represents number of households sampled and the proportion of households in that cluster relative to all households, respectively, and the proportions in the map are the proportion of study households by clusters (A); The probability distribution of the number of all household rooms compared to sleeping rooms (B); The probability distribution of the number of members in the household, with insert plot showing the proportion of smokers in all sampled households (C); Proportion of households using different sources of household energy (D); Proportion of households with indoor cooking (E); and proportion of households with indoor ventilation among households with indoor cooking (F).

**Supplementary Figure 2.**
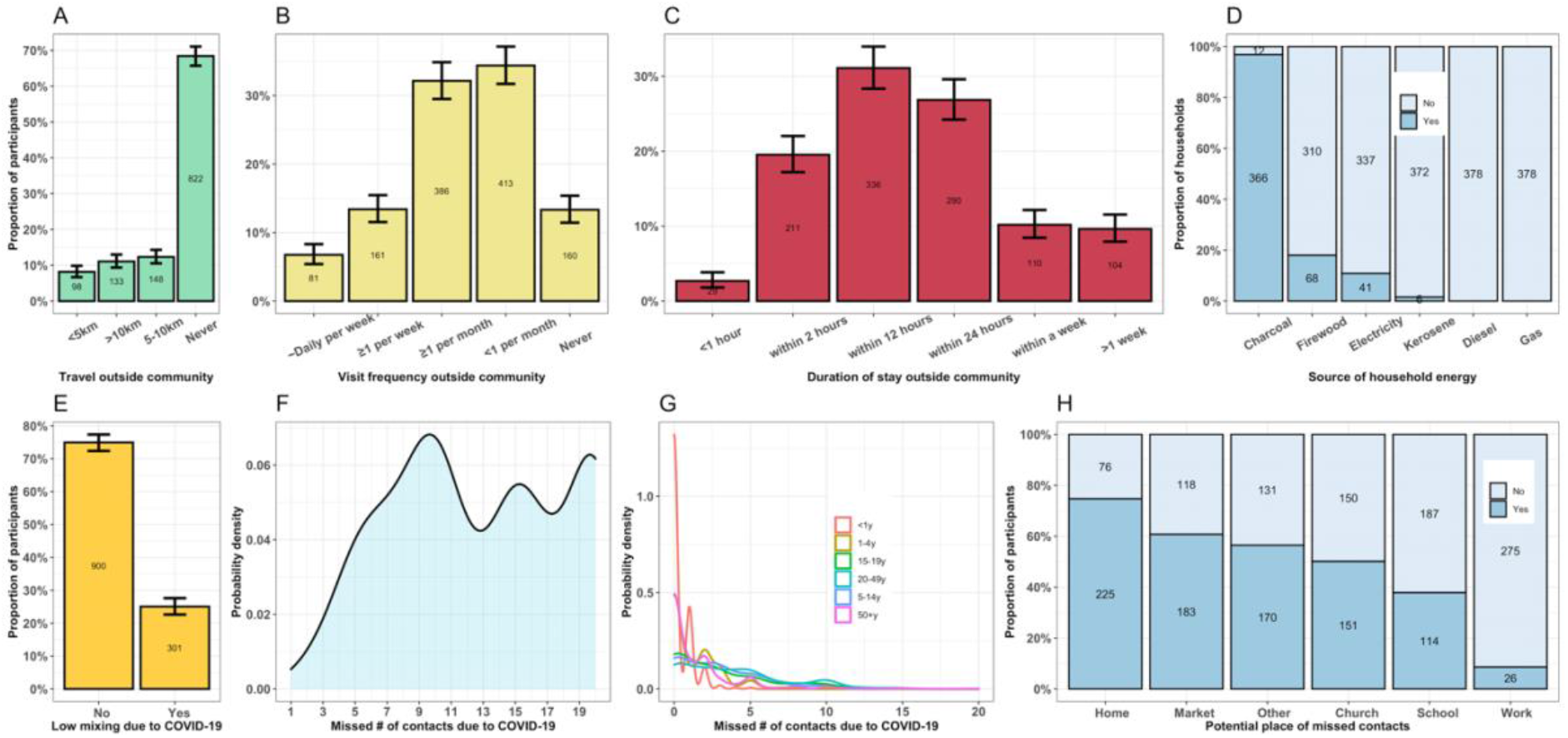
Travel history and COVID-19 related social behaviour. Proportion of participants: who travelled unique distances in the last 24 hours before initial study visit (A); with different visiting frequency outside Ndirande community (B); with different lengths of stay outside Ndirande community among those who make visits (C); using different means of travelling during visits outside Ndirande (D); and who reported having reduced number of contact events due to COVID-19 pandemic (E). The probability distribution of the number of missed contacts due to COVID-19 pandemic by overall (F) and age group (G). The proportion of hypothetical contacts and potential places where those contacts could have occurred in the absence of COVID-19 pandemic (H).

**Supplementary Figure 3.**
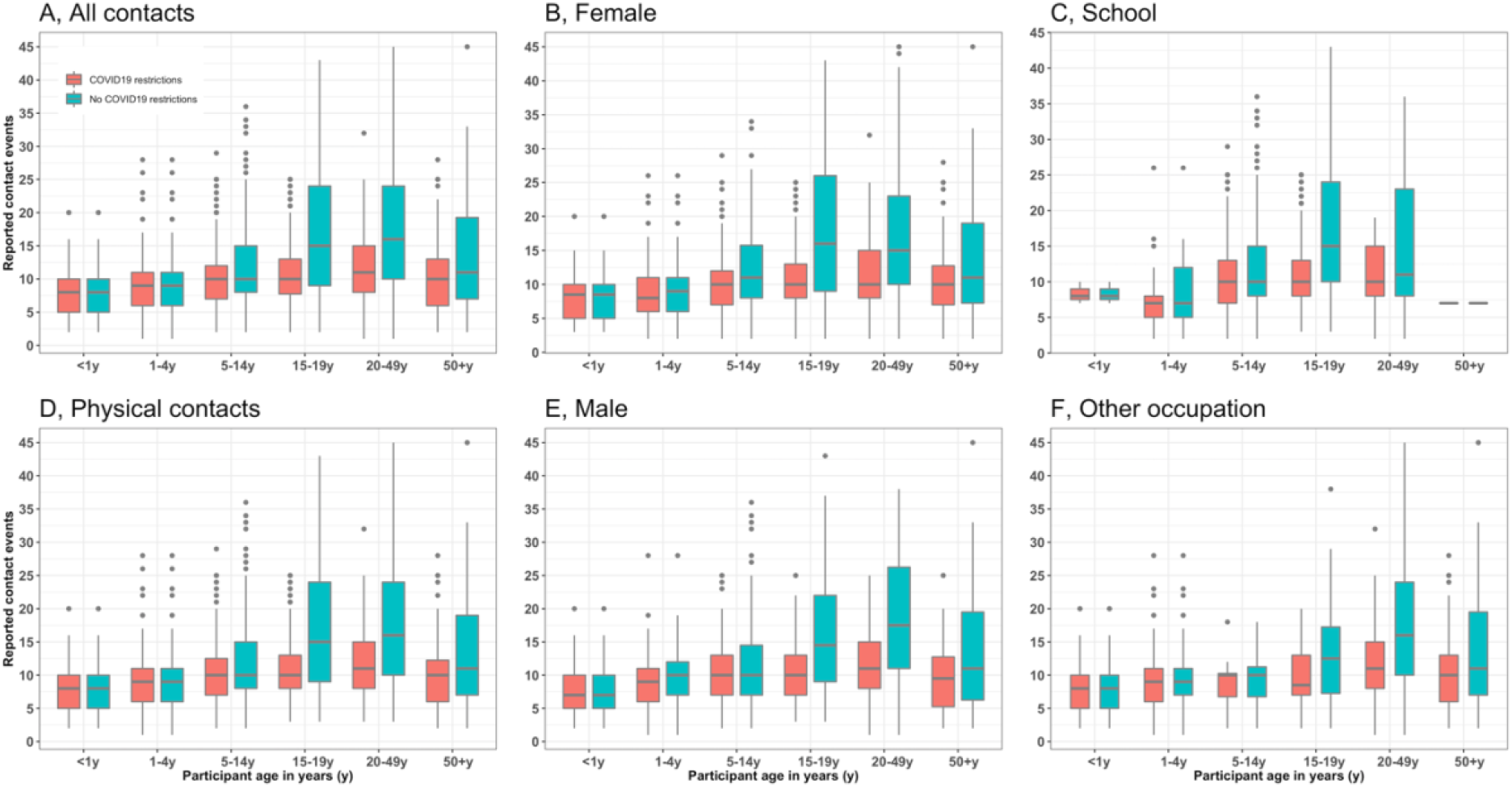
Distribution of contacts and contact events in those whose contact behaviour is affected or not affected by COVID-19 pandemic. The median, interquartile range, minimum and maximum number of contacts in those with and without restrictive social contact behaviour due to COVID-19, stratified by all mixing events (A); physical mixing events only (B); female sex (C); male sex (D); schoolers (E); and other occupation (F).

**Supplementary Figure 4.**
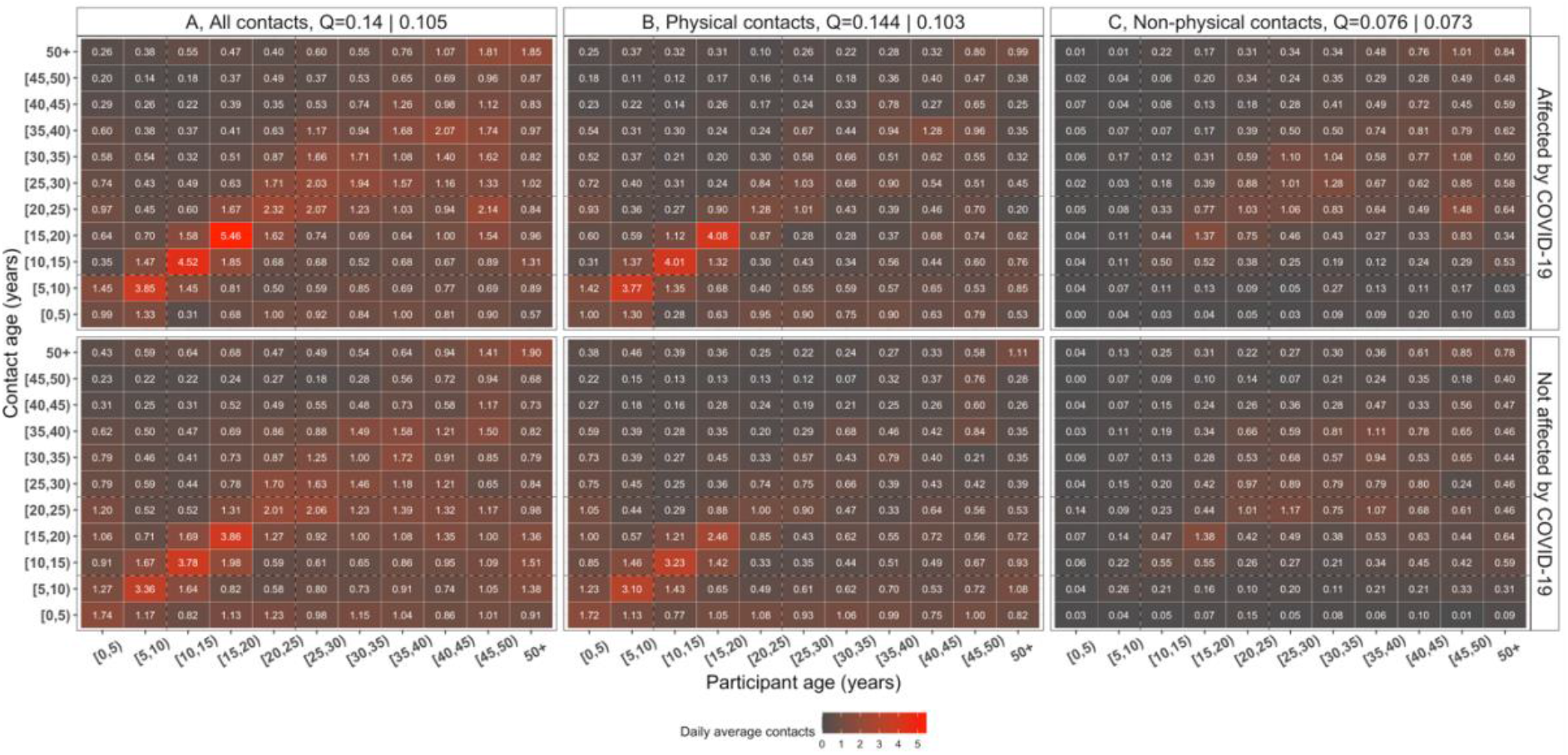
Social contact matrices by COVID-19 contacts behaviour change. The daily average rate of mixing between different age groups comparing participants who reported their social contact behaviour being affected and not affected by COVID-19 pandemic for all the contact events (A); physical contact events (B); and non-physical contact events (C). The number in each cell represents the daily mean number of contact events between two given age groups, corrected for mixing reciprocity between participants and contacts and weighted by day of the week. The assortativity index *Q* quantifies the weight of mixing between individuals of the same age groups.

**Supplementary Figure 5.**
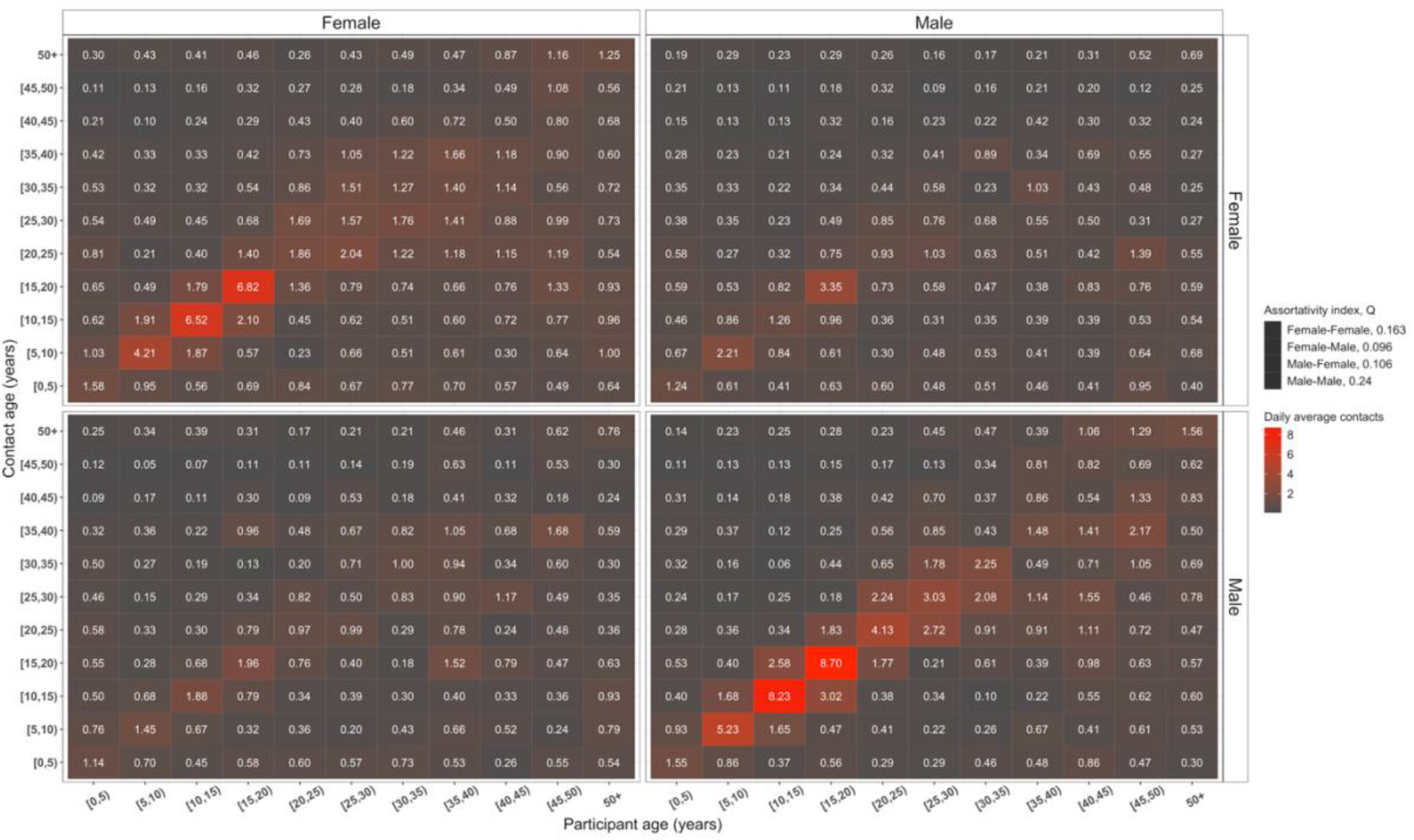
Social contact matrices by gender interactions. The daily average rate of mixing between different age groups comparing interactions between female participants and female contacts (top-left panel), male participants and female contacts (top-right panel), female participants and male contacts (bottom-left panel) and male participants and male contacts (bottom-right panel). The number in each cell represents the daily mean number of contact events between two given age groups, corrected for mixing reciprocity between participants and contacts and weighted by day of the week. The assortativity index *Q* quantifies the weight of mixing between individuals of the same age groups.

**Supplementary Table 1.** Ndirande community total population versus total sampled population and their respective proportions per age group.

|  |  |  |  |
| --- | --- | --- | --- |
| Age group, years old (y) | Population size<sup>†</sup> | Sample size | Proportion |
| <1y | 2414 | 65 | 2.7% |
| 1-4y | 10182 | 230 | 2.3% |
| 5-14y | 25780 | 265 | 1.0% |
| 15-19y | 11150 | 188 | 1.7% |
| 20-49y | 42417 | 301 | 0.7% |
| 50+y | 5388 | 152 | 2.8% |
| Total | 97,331 | 1,201 | 1.2% |
| † Data on the total population in each age group from previous census in Ndirande community (STRATAA study) (Thindwa et al., 2020) |  |  |  |

Thindwa, D., Farooq, Y.G., Shakya, M., Saha, N., Tonks, S., Anokwa, Y., Gordon, M.A., Hartung, C., Meiring, J.E., Pollard, A.J., Heyderman, R.S., The Strategic Typhoid alliance across Africa and Asia consortium, 2020. Electronic data capture for large scale typhoid surveillance, household contact tracing, and health utilisation survey: Strategic Typhoid Alliance across Africa and Asia. Wellcome Open Res 5, 66. https://doi.org/10.12688/wellcomeopenres.15811.2

## Supplementary Questionnaire

**PARTICIPANT SOCIAL MIXING QUESTIONNAIRE**

**STUDY NAME: SOCIAL MIXING PATTERNS (SOMIPA)**

**LOCATION: Blantyre**

**Section 1: Household and index identification**

|  |  |
| --- | --- |
| S1.1 SCALE household identification |  |
| S1.2 SCALE participant identification |  |
| S1.3 Community name |  |
| S1.4 Cluster number |  |
| S1.5 Household longitude |  |
| S1.6 Household latitude |  |
| S1.7 Individual age |  |
| S1.8 Individual sex | ☐ Female (0), ☐ Male (1) |

**Section 2: Household characteristics**

|  |  |
| --- | --- |
| S2.1 SOMIPA household ID (*scan barcode*) |  |
| S2.2 Interviewer ID |  |
| S2.3 How many separate rooms do you have? |  |
| S2.4 How many of these separate rooms are places to sleep? |  |
| S2.5 What is the source of fuel used inside the house?<br>(*select multiple answers if applicable*) | ☐ Gas (1/0) ☐ Charcoal (1/0)<br>☐ Electricity (1/0) ☐ Kerosene ethanol (1/0)<br>☐ Firewood (1/0) ☐ Diesel (1/0) |
| S2.6 Do you cook indoors? | ☐ Yes (1), ☐ No (0) |
| S2.7 If Yes, is ventilation present in the cooking space? | ☐ Yes (1), ☐ No (0), ☐ Don't know (99) |
| S2.8 When did your family settle in your current location? | ☐ <1 year (0) ☐ 5 years (5)<br>☐ 1 years (1) ☐ 6 years (6)<br>☐ 2 years (2) ☐ 7 years (7)<br>☐ 3 years (3). ☐ >7 years (8)<br>☐ 4 years (4). ☐ Don't know (99) |

**Section 3: Household composition**

|  |  |
| --- | --- |
| S3.1 Including yourself, how many individuals currently live in this household (do not include visitors)? | (enter "99" if you don't know) |
| S3.2 If there are any, how many household members have left to live somewhere else in the last 3 month? | (enter "99" if you don't know) |
| S3.3 If there are any, how many household members have died during the last 3 month? | (enter "99" if you don't know) |
| S3.4 Does anyone who currently lives in the household smoke cigarettes , chingambwe, or chamba? | ☐ Yes (1), ☐ No (0), ☐ Don't know (99) |

**Section 4: Participant Demographic**

|  |  |
| --- | --- |
| S4.1 SOMIPA household ID identification number<br>*scan a barcode sticker (assign new SOMIPA ID)* |  |
| S4.2 Household member number | (*member number 1 is for index participant*) |
| S4.3 Who is a respondent to this interview? | Self (0),<br>Parent/Guardian (1) |
| S4.4 Do you know your exact date of birth? | Yes (1) *go to S4.5*<br>No (0) *go to S4.6* |
| S4.5 What is your exact date of birth?<br>*check health passport book to confirm if available* | d<br>d<br>m<br>m<br>y<br>y<br>y<br>y |
| S4.6 What is your age? |  |
| S4.7 Is your age in months or years? | Months (0),<br>Years (1) |
| S4.8 Is your age exact or approximated? | Exact (0),<br>Approximated (1) |
| S4.9 How was age estimated? | Local event calendar (1)<br>Proxy age of the other person (2)<br>Guess (3) |
| S4.10 What is your sex? | Female (0),<br>Male (1) |
| S4.11 What is your primary occupation or daily activity? | Pre-school child (1)<br>School/college/university student (2)<br>Office worker (3)<br>Shop worker (4)<br>Manual worker (5)<br>Agriculture (6)<br>Market salesman (7)<br>Household (8)<br>Unemployed (9)<br>Retired (10)<br>Other (11) _____<br>Refuse to answer (88)<br>Don’t know (99) |
| S4.12 What is your highest level of education attended?<br>*only for adults >=18 years* | No education (1)<br>Primary school (2)<br>Secondary school (3)<br>College/university (4) |

**Section 5: Travel & COVID-19**

|  |  |
| --- | --- |
| S5.1 How furthest did you travel outside your community in the last 24 hours? | ☐ <5 km away, <1 hour walk (1)<br>☐ 5-10 km away, 1-2 hours walk (2)<br>☐ >10 km away (3), > 2 hours walk<br>☐ Never (4)<br>☐ Refuse to respond (88) |
| S5.2 How often do you visit places outside your community? | ☐ Most days of the week (1)<br>☐ At least once per week (2)<br>☐ At least once per month (3)<br>☐ Less than once per month (4)<br>☐ Never (5)<br>☐ Refuse to respond (88) |
| S5.3 When you go to places outside your community, how much time do you spend there on a maximum? | ☐ <1 hour (1)<br>☐ 1-2 hours (2)<br>☐ Half a day (3)<br>☐ A whole day (4)<br>☐ One week (5)<br>☐ More than one week (6)<br>☐ Don't know (99)<br>☐ Refuse to answer (88) |
| S5.4 What is your main means of transport when travelling?<br>(multiple ticks allowed) | ☐ Walk on foot (1)<br>☐ Bicycle (2)<br>☐ Personal car (3)<br>☐ Public transport (4)<br>☐ Carried on the back (5)<br>☐ Don't know (99)<br>☐ Refuse to answer (88) |
| S5.5 Are your encounters less now due to COVID-19? | ☐ Yes (1), ☐ No (0), ☐ Don't know (99)<br>(if Yes, go to S5.6) |
| S5.6 How many additional people would you have encountered from yesterday if there was no COVID-19? |  |
| S5.7 Where would those encounters/contacts have happened?<br>(multiple ticks allowed) | ☐ Home or another house (1)<br>☐ Work (2)<br>☐ School (3)<br>☐ Church/Mosque (4)<br>☐ Shop/Grocery/Market (5)<br>☐ Leisure/Water source/garden/Other (6) |
| S5.8 What age groups would those contacts have been?<br>(indicate how many contacts in each age group) | Infants, less than 1 year old<br>Pre-school children, 1-5 years old<br>Primary school children, 6-15 years old<br>Secondary school children 16-19 years old<br>Adults, 20-49 years old<br>Elderly, at least 50 years old |

|  | Type of Place |  |  |  |  |  |  |  |  |  |  |  |  | Location |  |
| --- | --- | --- | --- | --- | --- | --- | --- | --- | --- | --- | --- | --- | --- | --- | --- |
| Place ID | Home | Another house | Work | School | Mosque/church | Transport | Leisure | Shop/ grocery | Market | Water source | Garden | Other | Name other | Inside the community | Outside the community |
| 1 | ☐ | ☐ | ☐ | ☐ | ☐ | ☐ | ☐ | ☐ | ☐ | ☐ | ☐ | ☐ |  | ☐ | ☐ |
| 2 | ☐ | ☐ | ☐ | ☐ | ☐ | ☐ | ☐ | ☐ | ☐ | ☐ | ☐ | ☐ |  | ☐ | ☐ |
| 3 | ☐ | ☐ | ☐ | ☐ | ☐ | ☐ | ☐ | ☐ | ☐ | ☐ | ☐ | ☐ |  | ☐ | ☐ |
| 4 | ☐ | ☐ | ☐ | ☐ | ☐ | ☐ | ☐ | ☐ | ☐ | ☐ | ☐ | ☐ |  | ☐ | ☐ |

**Section 6: Contact information**

*I will finally ask you to recall who you were in contact with between now and yesterday? I will ask this for each of places you listed previously. Now I am only interested in your direct contacts. These are individuals with who you spent ≥5 mins, and with whom you spoke ≥3 words. For each of contact. I would like to know their age in years, sex, your relationship to him/her, where the contact happened, how often you contact this person in general, and the total time spent with this person in that place. I would also like to know whether contact was physical or nonphysical. Nonphysical contact happens when you haven’t touched the person. Physical contact includes hand shaking, embracing, kissing, and sharing a bike, and also sharing a glass or other utensils passed directly from mouth to mouth. I only ask for contact initials to help remember the contact, but will delete the information at the end of the interview. If you had contact with same person in multiple settings, I will record these multiple times, once for each setting. If you don’t know their exact age, you can take a best guess.*

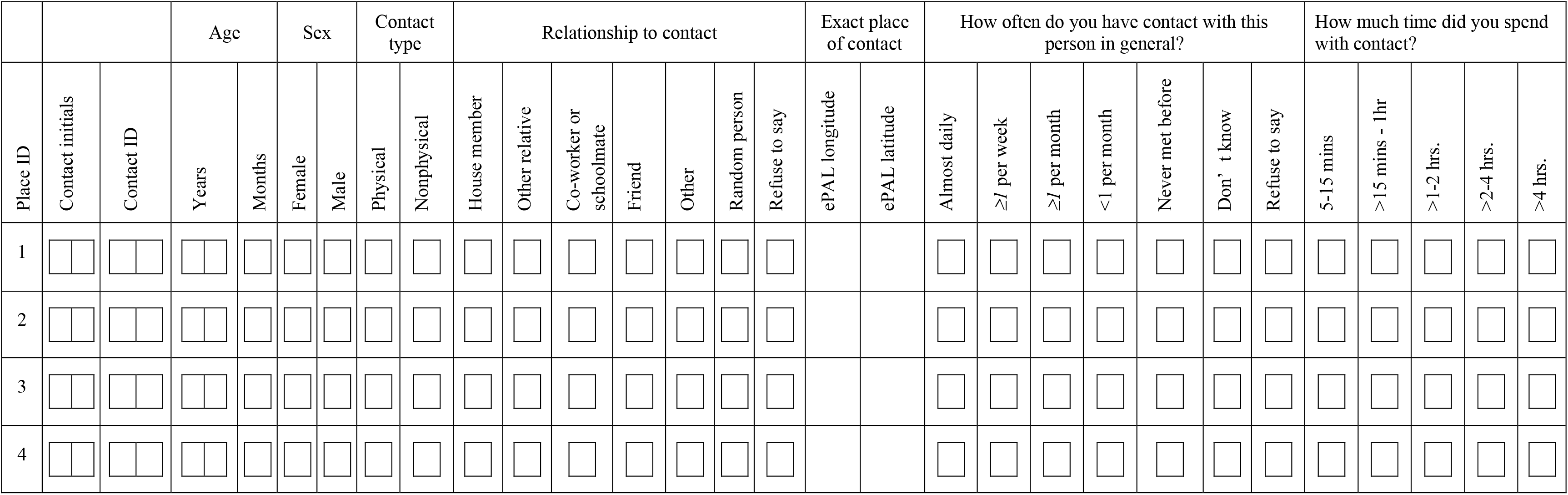

|  |  |  | Age |  | Sex |  | Contact type |  | Relationship to contact |  |  |  |  |  |  | Exact place of contact |  | How often do you have contact with this person in general? |  |  |  |  |  |  | How much time did you spend with contact? |  |  |  |  |
| --- | --- | --- | --- | --- | --- | --- | --- | --- | --- | --- | --- | --- | --- | --- | --- | --- | --- | --- | --- | --- | --- | --- | --- | --- | --- | --- | --- | --- | --- |
| Place ID | Contact initials | Contact ID | Years | Months | Female | Male | Physical | Nonphysical | House member | Other relative | Co-worker or schoolmate | Friend | Other | Random person | Refuse to say | ePAL longitude | ePAL latitude | Almost daily | ≥1 per week | ≥1 per month | <1 per month | Never met before | Don't know | Refuse to say | 5-15 mins | >15 mins - 1hr | >1-2 hrs. | >2-4 hrs. | >4 hrs. |
| 1 |  |  |  |  |  |  |  |  |  |  |  |  |  |  |  |  |  |  |  |  |  |  |  |  |  |  |  |  |  |
| 2 |  |  |  |  |  |  |  |  |  |  |  |  |  |  |  |  |  |  |  |  |  |  |  |  |  |  |  |  |  |
| 3 |  |  |  |  |  |  |  |  |  |  |  |  |  |  |  |  |  |  |  |  |  |  |  |  |  |  |  |  |  |
| 4 |  |  |  |  |  |  |  |  |  |  |  |  |  |  |  |  |  |  |  |  |  |  |  |  |  |  |  |  |  |

## Notes

### Competing Interest Statement

The authors have declared no competing interest.

### Author Declarations

Ethical approval was granted by the College of Medicine Research Ethics Committee, University of Malawi (P.01/21/3244), and the London School of Hygiene and Tropical Medicine Research Ethics Committee (#22913). Informed consent was obtained from each participant aged >16 years, or from the parent or guardian of each individual aged <6 years, or from each participant aged 6-15 years with addition informed assent.

